# A safe protocol to identify low risk patients with COVID-19 pneumonia for outpatient management

**DOI:** 10.1101/2020.12.15.20229286

**Authors:** Francisco Javier Teigell Muñoz, Elena García-Guijarro, Paula García-Domingo, Guadalupe Pérez-Nieto, Fernando Roque Rojas, María García-Peña, María Antonia Nieto Gallo, Jose Antonio Melero Bermejo, Maria Teresa de Guzman García-Monge, Juan José Granizo

## Abstract

**Background:** The coronavirus disease 2019 (COVID-19) outbreak has made necessary to rationalize health-care resources, but there are no published data to this moment regarding ambulatory management of patients with COVID-19 pneumonia.

**Objective:** Evaluate the results of a protocol for ambulatory management of patients with COVID-19 pneumonia according to the rate of readmissions, admission into the Intensive Care Unit (ICU) and deaths. Identify unfavorable prognostic factors that increase the risk of readmission, ICU admission and/or death.

**Methods:** Prospective cohort study of patients with COVID-19 pneumonia discharged from the emergency ward of Infanta Cristina Hospital (Madrid, Spain), that met the criteria of the hospital protocol for outpatient management. We describe outcomes of those patients and compare those who needed readmission versus those we did not. We use logistic regression to explore factors associated with readmissions.

**Findings:** 314 patients were included, of which 20 (6.4%) needed readmission, 3 (1%) developed severe respiratory failure, and none needed ICU admission nor died. 29.9% of patients had any one comorbidity. Hypertension, leukopenia, lymphocytopenia, increased lactate dehydrogenase (LDH), increased aminotransferases were associated to a higher risk of readmission. A clinical course of 10 days or longer, and an absolute eosinophil count over 200/µL were associated with a lower risk. After multivariate analysis, only hypertension (OR 4.99, CI 1.54-16.02), temperature over 38°C in the emergency ward (OR 9.03, CI: 1.89-45.77), leukopenia (OR 4.92, CI 1.42-17.11) and increased LDH (OR 6.62, CI 2.82-19.26) remained significantly associated to readmission.

**Conclusion:** Outpatient management of patients with low-risk COVID-19 pneumonia is safe, if adequately selected. The protocol presented here has allowed avoiding 30% of the admissions for COVID-19 pneumonia in our hospital, with a very low readmission rate and non-existing mortality.

**Funding:** The authors received no specific funding for this work.

## INTRODUCTION

Spain has been one of the countries most severely hit by the worldwide SARS-CoV-2 pandemic, with over 230,000 confirmed cases by May 31^th^ 2020 and approximately 28,000 deaths^1^. When COVID-19 arrived in Spain, little was known about its course and development. Although most patients only presented with mild episodes, many developed pneumonia and some of them developed severe respiratory failure and, eventually, death. As it was especially difficult to identify the patients at higher risk of unfavorable evolution, the initial clinical strategy was to admit all patients diagnosed with COVID-19 and pneumonia for treatment and close monitoring.

However, the great accumulated incidence of pneumonia cases in a short time span, showing an exponential growth of the amount of hospital admissions, led not only to a shortening in material and human resources, but also to a high risk of collapse for the health system, which could itself lead to a decrease in the quality of patient care and hence to an increase in mortality.

It became necessary to create a strategy to rationalize health-care resources and prioritize inpatient attention to those in greater need. Within the Section of Emergency Medicine and Internal Medicine at the Infanta Cristina Hospital (ICH) in Madrid, Spain, we developed a protocol in order to select those patients which would truly benefit from inpatient care and those who could be cared for via outpatient telephonic follow-up, based on established criteria.

In this study, we describe the protocol of ambulatory care for patients with low risk COVID-19 pneumonia and evaluate its results in our setting during the current pandemic.

## METHODS

### Study design and sample

A cohort of patients discharged between March 17^th^ and April 25^th^ 2020 from the emergency ward of the ICH with clinical and radiological diagnosis of COVID-19 pneumonia was prospectively evaluated.

Among all patients managed as outpatients, only those who met the discharge criteria established in the hospital protocol were included. Said criteria (Table 1) were elaborated after reviewing the literature available at the moment, emphasizing the work of and Fei Zhou *et al*,^2^ and Chaomin Wu *et al*,^3^ where they describe both favorable and unfavorable prognostic factors in COVID-19 patients. According to said protocol, a chest radiograph was performed on all patients with fever and/or other symptoms suggesting COVID-19. Those with no pathological findings in the radiograph were discharged whenever their clinical status allowed for it. In radiologically dubious cases, chest CT or lung ultrasound were performed. Blood tests including blood cell count, biochemical analysis and coagulation study were performed to all patients with radiological image compatible with COVID-19 pneumonia; in case of hypoxemia, blood gas analysis was also performed. The protocol also recommended performing a simplified walking-test in order to assess lung capacity. Said test consisted in having the patient walk fast 30 meters; it was considered positive whenever oxygen saturation, as measured by pulse oximetry, decreased by 5 percent or more. RT-PCR for SARS-CoV-2 was performed via nasopharyngeal swab, according to the instructions from the official health authority at the time of each evaluation. Patients with suspected COVID-19 pneumonia could be discharged whenever they did not meet major admission criteria (Table 1). Admission was to be considered in cases with multiple coexisting minor criteria. However, and due to the absence of strong evidence, the ultimate decision was always left to the judgment of the physician in charge. The telephone number of the patient was registered upon discharge and the case was reported to the follow-up team in order to continue ambulatory surveillance. All patients were discharged with home therapy, according to the protocol of the hospital valid at the time of discharge.

**Table 1.** Discharge criteria according to the protocol of the Infanta Cristina Hospital

| Infanta Cristina Hospital's discharge criteria<br>(no major criteria should be met, some minor criteria are allowed) |  |
| --- | --- |
| Major criteria | Minor criteria |
| Age > 70 years | Age 50-70 years |
| SaO<sub>2</sub> < 95% and/or positive walking-test | Comorbidities* (1 point each) |
| Respiratory rate > 30 | Increased aminotransferases |
| Bronchospasm | Increased LDH |
| Radiological affection > 50% |  |
| CPR > 100 mg/L | CPR 50-100 mg/L |
| D-dimer 1,000 ng/mL | D-dimer 500-1,000 ng/mL |
| Lymphocytes < 800/μL | Lymphocytes 800-1,200/μL |
| CURB-65 score ≥ 2 | |

SaO<sub>2</sub>: baseline oxygen saturation; LDH: lactate-dehydrogenase; CPR: C-reactive protein; CURB65: confusion, urea, respiratory rate, low blood pressure, and age >65 years.
\*Comorbidities: Persistent asthma, chronic obstructive pulmonary disease, obstructive sleep apnea-hypopnea syndrome, ischemic heart disease, active cancer, kidney failure, liver failure, hypertension, diabetes mellitus, obesity, immunosuppression.

The telephone follow-up started the day after discharge, which could be daily or once every other day, according to the patient’s status. In each call, a basic questionnaire was performed, which included questions about the presence of fever, dyspnea, cough, chest pain, digestive symptoms and drug tolerability. The importance of isolation measures was also emphasized during the call. Patients with non-relenting fever despite medication, fever persisting longer than 3 days after the start of follow-up, resting or minimal-exertion dyspnea, inability to feed or limiting asthenia, were referred back to the emergency ward for on-site evaluation, complementary work-up or, eventually, inpatient admission. During the study period, some digital pulse oximeters became available, and they were lent to the patients at highest risk, which allowed for a more adequate surveillance of their respiratory status.

Those patients that did not meet the protocol criteria but were also discharged (e.g. those meeting major admission criteria) were excluded from analysis, as were those for which the telephone follow-up was deemed unfeasible because of mental illness, language barrier, etc. Pregnant patients have also been excluded, as they were followed by the Section of Gynecology and Obstetrics of our institution. Patients with intermediate to low clinical suspicion and negative RT-PCR were also excluded.

### Study objectives

The main goal was to evaluate the results of the protocol for ambulatory management according to the rate of readmissions due to complications or worsening, the rate of admission into the Intensive Care Unit (ICU) and the rate of deaths. Secondary objective was to identify unfavorable prognostic factors that increase the risk of readmission, ICU admission and/or death.

### Statistical analysis

IBM SPSS 16.0 was used for analysis. Quantitative variables are displayed as median, interquartile range (IQR) and eventually range; categorical variables are displayed as absolute and relative (%) frequency. For comparative analysis, both the χ^2^ and Fisher’s exact test were used in the case of categorical variables, and Mann-Whitney’s test for non-categorical quantitative variables. In order to explore the risk factors associated to readmission, both bivariate and multivariate logistic regression models were used. After bivariate analysis, considering the low rate of readmission (20 in total), only 4 variables were used in the multivariate analysis. Only those differences with an associated bilateral alpha error lower than 0.05 were considered significant.

## RESULTS

Clinical and demographical features of the sample are shown in Table 2. The age median was 45 years (IQR 40-53), and range was 18-70. 52.5% were female. 29.9% of the patients had any one comorbidity (median 0, IQR 0-1), and 10% had 2 or more simultaneous comorbidities (range 0-5). The most common ones was hypertension (14.3%), followed by diabetes mellitus (DM) and obesity (5.4% each), and persistent asthma (4.8%). No patient had kidney or liver failure. The median time from the initial symptoms was 7 days, and 37.7% of the patients presented on the 10^th^ day of disease progression or later. 71.0% showed a normal pulmonary auscultation, and only 3.8% of the patients had a temperature equal to or higher than 38°C at the time of evaluation.

**Table 2.** Demographic and clinical features

|  | Total | Readmission | No readmission | p value |
| --- | --- | --- | --- | --- |
|  | n = 314 | n = 20 | n = 294 |  |
| Age (years) | 45.0 (40.0-53.0) | 46.5 (42.3-50.8) | 45 (39-53) | 0.501 |
| 50-70 | 107 (34.1%) | 6 (30%) | 101 (34.4%) | 0.69 |
| Sex |  |  |  |  |
| Female | 165 (52.5%) | 8 (40%) | 157 (53.4%) | 0.24 |
| Male | 149 (47.5%) | 12 (60%) | 137 (46.6%) |  |
| Active smoking | 21/124 (6.7%) | 1/9 (11.1%) | 20/115 (17.4%) | 0.38 |
| Comorbidities |  |  |  |  |
| Number of comorbidities per patient | 0 (0-1) | 0 (0-1) | 0 (0-1) | 0.369 |
| Presence of at least one comorbidity | 94 (29.9%) | 8 (40%) | 86 (29.3%) | 0.144 |
| Persistent asthma | 15 (4.8%) | 0 (0%) | 15 (5.1%) | 0.36 |
| COPD | 1 (0.3%) | 0 (0%) | 1 (0.3%) | - |
| OSAHS | 4 (1.3%) | 0 (0%) | 4 (1.3%) | - |
| Ischemic heart disease | 5 (1.6%) | 1 (5%) | 4 (1.4%) | 0.282 |
| Active cancer | 1 (0.3%) | 0 (0%) | 1 (0.3%) | - |
| Kidney/liver failure | 0 (0%) | - | - |  |
| Hypertension | 45 (14.3%) | 6 (30%) | 39 (13.3%) | 0.039 |
| Use of ACE-inhibitors or ARBs | 35 (11.2%) | 4 (20%) | 31 (10.5%) | 0.26 |
| Diabetes mellitus | 17 (5.4%) | 3 (15%) | 14 (4.8%) | 0.085 |
| Obesity | 17 (5.4%) | 3 (15%) | 14 (4.8%) | 0.085 |
| Use of immune suppressants | 10 (3.2%) | 0 (0%) | 10 (3.4%) | 0.512 |
| Time since symptom start | 7.0 (5-12) | 7 (4.3-7) | 8 (5-12) | 0.009 |
| <10 days | 193 (62.3%) | 20 (100%) | 177 (59.7%) |  |
| ≥10 days | 117 (37.7%) | 0 (0) | 117 (40.3%) | <0.001 |
| Fever (told by the patient) | 238 (75.8%) | 18 (90%) | 220 (75%) | 0.17 |
| Cough | 246 (78.3%) | 16 (80%) | 230 (78.5%) | 0.87 |
| Dyspnea | 190 (60.5%) | 12 (60%) | 178 (60.8%) | 0.95 |
| Diarrhea or vomiting | 100 (31.8%) | 6 (30%) | 94 (32.2%) | 0.84 |
| Arterial oxygen saturation in the emergency ward | 98 (96-98) | 97.1 (96-98) | 97.6 (96-98) | 0.18 |
| Heart rate in the emergency ward (bpm) | 89 (80-200) | 90 (85-104) | 91.5 (80-99) | 0.565 |
| Temperature in the emergency ward (°C) | 36.6 (36.4-36.9) | 36.6 (36.4-37.1) | 36.6 (36.4-36.9) | 0.634 |
| ≥38°C | 11/314 (3.5%) | 4 (20%) | 7 (2.4%) | 0.003 |
| Abnormal lung auscultation | 91 (29%) | 8 (40%) | 83 (28.3%) | 0.27 |

COPD: chronic obstructive pulmonary disease; OSHAS: Obstructive sleep apnea-hypopnea syndrome; ACE: angiotensin-converting enzyme; ARB: angiotensin receptor blockers.

Laboratory tests are available for all but one patient (Table 3). Among the most commonly seen abnormalities were elevated aminotransferases (38.5%), elevated D-dimer (27.8%), low lymphocyte count (25.5%) or low total white blood cell count (8.6%). All patients had a CURB-65 score of 0 to 1 points (94.6 and 5.4% respectively). The statistical distribution of radiological pulmonary abnormalities is shown in Table 3. The most common infiltrate pattern was a bilateral affection under 50% of the lung fields (54.8%), followed by unilobar affection (33.4%) and unilateral multilobar affection (10.5%). Three patients had a normal chest radiograph, and the diagnosis of pneumonia was made via lung ultrasound or chest tomography. RT-PCR for SARS-CoV-2 was performed on 138 (43.9%) patients; 93 were positive (67.4% of the performed PCR tests), 40 were negative (29.0%) and 5 gave indeterminate results (3.6%). The patients included in the ambulatory management program had a median of two minor criteria (IQR 1-3; range 0-8). Only 11.1% of patients had four or more minor criteria. Among them, 15.6% required readmission.

**Table 3.** Laboratory, imaging and microbiology test results.

|  | Total | Readmission | No readmission | p value |
| --- | --- | --- | --- | --- |
| **Laboratory findings** | n = 313 | n = 19 | n = 294 |  |
| Leukocytes (/μL) | 6390 (5132-7960) | 4920 (3960-6300) | 6520 (5220-8110) | 0.001 |
| <4000 | 27 (8.6%) | 5 (26.3%) | 22 (7.4%) | 0.004 |
| Lymphocytes (/μL) | 1599 (1100-2000) | 1200 (1093-1700) | 1600 (1200-2000) | 0.041 |
| 800-1200 | 80 (25.5%) | 9 (47.4%) | 71 (24.1%) | 0.025 |
| >=1200 | 233 (74.5%) |  |  |  |
| Eosinophils (/μL) | 100 (0-100) | 0 (0-0) | 100 (0-100) | <0.001 |
| >200 | 28/301 (9.3%) | 0 (0%) | 28/282 (9.9%) | 0.234 |
| Platelets (x10<sup>3</sup>/μL) | 226 (186-283) | 163 (133-189) | 231 (190-286) | <0.001 |
| <100 | 1 (0.3%) | 1 (5.3%) | 0 (0%) | 0.06 |
| Lactate-dehydrogenase (U/L) | 197 (166-235) | 253 (234-287) | 193 (164-228) | <0.001 |
| >250 | 46/270 (16.5%) | 9/18 (50%) | 37/261 (14.2%) | <0.001 |
| Creatinine (mg/dL) | 0.83 (0.7-0.98) | 0.98 (0.81-1.15) | 0.82 (0.70-0.97) | 0.005 |
| ≥1.3 mg/dL | 5 (1.64%) | 1 (5.3%) | 4 (1.4%) | 0.227 |
| Aspartate-aminotransferase (U/L) | 29 (21-37.25) | 38 (33-48) | 28 (21-37) | 0.004 |
| Alanine-aminotransferase (U/L) | 38 (28-55) | 46 (36.5-60.5) | 38 (28-55) | 0.299 |
| AST> 40 y/o ALT > 50 | 121 (38.5%) | 12 (60%) | 109 (37.1%) | 0.042 |
| C-reactive protein (mg/L) | 18.6 (4.5-35) | 29.3 (17.3-29.3) | 16.8 (4.3-34.3) | 0.012 |
| <50 | 264 (84.4%) |  |  |  |
| 50-100 | 49 (15.6%) | 5 (26.3%) | 44 (15.5%) | 0.207 |
| Prothrombin activity (%) | 92 (84-98) | 98 (89-102) | 91 (84-98) | 0.175 |
| D-dimer (ng/mL) | 360 (250-530) | 365 (267-435) | 360 (250-530) | 0.901 |
| <500 | 226 (72.2%) |  |  |  |
| 500-1000 | 87 (27.8%) | 4 (21%) | 83 (28.4%) | 0.569 |
| **CURB-65** | 0 (0-0) | 0 (0-0) | 0 (0-0) |  |
| =0 | 297 (94.6%) | 18 (94.7%) | 279 (94.7%) |  |
| =1 | 17 (5.4%) | 1 (5.3%) | 16 (5.4%) | - |
| **Radiological findings** |  | n = 20 | n = 294 | 0.223 |
| Normal radiograph | 3 (1%) | 3 (1%) | 0 (0%) |  |
| Unilobar affection | 105 (33.4%) | 4 (20%) | 101 (34.4%) | 0.188 |
| Unilateral multilobar affection | 33 (10.5%) | 5 (25%) | 28 (9.5%) |  |
| Bilateral affection < 50% | 172 (54.8%) | 11 (55%) | 161 (54.8%) |  |
| **Virological diagnosis** |  | n = 20 | n = 294 |  |
| RT-PCR SARS-CoV-2 performed | 138/314 (43.9%) | 15 | 123 |  |
| Positive (% of performed) | 93 (67.4%) | 14 (93.3%) | 79 (64%) | 0.075 |
| Negative (% of performed) | 40 (29%) |  |  |  |
| Inhibited or indeterminate (% of performed) | 5 (3.6%) |  |  |  |
| RT-PCR SARS-CoV-2 not performed | 176 (56.1%) |  |  |  |

\* In those cases, the diagnosis was made via chest CT or lung ultrasound.

Table 4 shows the therapy prescribed upon discharge. It shows also the data of the follow-up and the development of the patients. The telephone follow-up period had a median of 5 days starting from the day of discharge (IQR 4-6, range 1-11), with a median of 3 telephone calls per patient (IQR 3-4, range 1-10). Depending on the availability in each moment, in the end 15.6% of patients were given a portable pulse oximeter to facilitate their monitoring. 62 patients (19.7%) were reevaluated in the emergency ward at least one second time. Half of them (9.9%) were referred because of medical criteria (almost all of them because of persistent fever or dyspnea), whereas the other half (9.9%) presented by their own choice. 42 of those patients were newly discharged, so 20 patients (6.4%) ultimately needed inpatient hospital admission. The most common reason for readmission (50%) was respiratory worsening with development of hypoxemia (SatO2 < 95%), with one only case of respiratory failure (SatO2 88%). One patient was readmitted with a diagnosis of segmental pulmonary embolism. 60% of readmissions presented again by their own choice. All patients that were readmitted did so within the first 6 days of follow-up (median 3, IQR 2-4, range 1-6). During admission, three patients developed severe respiratory failure. No patients needed ICU admission or died.

**Table 4.**
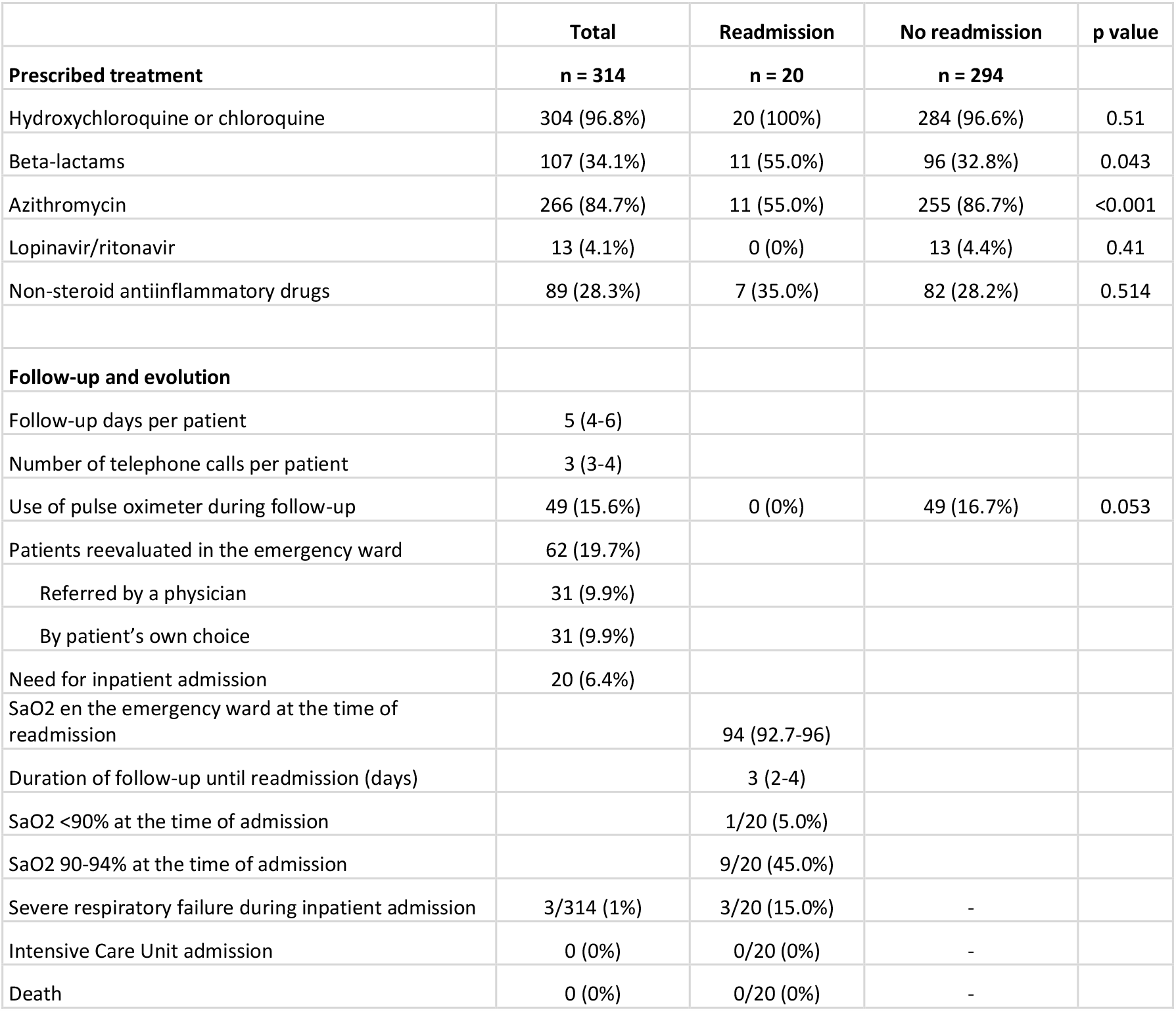
Prescribed treatment, follow-up and evolution.

|  | **Total** | **Readmission** | **No readmission** | **p value** |
| --- | --- | --- | --- | --- |
| **Prescribed treatment** | **n = 314** | **n = 20** | **n = 294** |  |
| Hydroxychloroquine or chloroquine | 304 (96.8%) | 20 (100%) | 284 (96.6%) | 0.51 |
| Beta-lactams | 107 (34.1%) | 11 (55.0%) | 96 (32.8%) | 0.043 |
| Azithromycin | 266 (84.7%) | 11 (55.0%) | 255 (86.7%) | <0.001 |
| Lopinavir/ritonavir | 13 (4.1%) | 0 (0%) | 13 (4.4%) | 0.41 |
| Non-steroid antiinflammatory drugs | 89 (28.3%) | 7 (35.0%) | 82 (28.2%) | 0.514 |
| **Follow-up and evolution** |  |  |  |  |
| Follow-up days per patient | 5 (4-6) |  |  |  |
| Number of telephone calls per patient | 3 (3-4) |  |  |  |
| Use of pulse oximeter during follow-up | 49 (15.6%) | 0 (0%) | 49 (16.7%) | 0.053 |
| Patients reevaluated in the emergency ward | 62 (19.7%) |  |  |  |
| Referred by a physician | 31 (9.9%) |  |  |  |
| By patient's own choice | 31 (9.9%) |  |  |  |
| Need for inpatient admission | 20 (6.4%) |  |  |  |
| SaO2 en the emergency ward at the time of readmission |  | 94 (92.7-96) |  |  |
| Duration of follow-up until readmission (days) |  | 3 (2-4) |  |  |
| SaO2 <90% at the time of admission |  | 1/20 (5.0%) |  |  |
| SaO2 90-94% at the time of admission |  | 9/20 (45.0%) |  |  |
| Severe respiratory failure during inpatient admission | 3/314 (1%) | 3/20 (15.0%) | - |  |
| Intensive Care Unit admission | 0 (0%) | 0/20 (0%) | - |  |
| Death | 0 (0%) | 0/20 (0%) | - |  |

### Secondary objectives

The patients requiring readmission were compared with the ones that did not require it in order to detect significant differences. As shown in Table 2, there were no differences regarding age or sex. Among the patients that required readmission, there was a higher proportion of hypertension (30.0% vs. 13.3%, p 0.039). The frequency of diabetes and obesity was also higher, but did not reach statistical significance (15% vs. 4.8% in both cases, p 0.085). There was no difference regarding other comorbidities, neither in the total amount of comorbidities per patient. Referred symptoms were similarly distributed in both groups, but a temperature equal or higher 38°C measured within the emergency ward was more frequent among the patients that did require readmission (20.0% vs. 2.4%, p 0.003). The readmitted patients showed a significantly shorter time period since the initial symptoms (7.0 vs. 8.0 days; p 0.009). Not one patient with 10 or days of disease progression at the time of presentation was readmitted.

The patients that required readmission showed significantly lower total white blood cell, lymphocyte, eosinophil and platelet counts, and subsequently a higher rate of leukopenia, lymphocytopenia and thrombocytopenia (Table 3). There were no readmissions among the 28 patients with more than 200 eosinophils per microliter. Readmitted patients had higher levels of LDH, C reactive protein, creatinine and liver enzymes. There were no differences in D-dimer or prothrombin activity.

No significant differences were detected in radiological patterns, although a lower rate of unilobar pneumonia was observed in the patients that were readmitted (Table 3). There was also a higher proportion of patients with positive RT-PCR in that group, but it did not reach statistical signification.

The readmitted patients received beta-lactams significantly more (55.0 vs. 32.8%, p 0.043) and azithromycin significantly less (55.0 vs. 86.7%, p <0.001). There were no differences between groups regarding hydroxychloroquine or chloroquine, lopinavir/ritonavir or NSAIDs.

In the bivariate analysis, hypertension, leukopenia, lymphocytopenia, increased LDH, increased aminotransferases and use of beta-lactams were associated to a higher risk of readmission (Table 5). On the contrary, a clinical course of 10 days or longer, an absolute eosinophil count over 200/µL and use of azithromycin were associated with a lower risk. However, after multivariate analysis only hypertension (OR 4.99, CI 1.54-16.02), temperature over 38°C in the emergency ward (OR 9.03, CI: 1.89-45.77), leukopenia (OR 4.92, CI 1.42-17.11) and increased LDH (OR 6.62, CI 2.82-19.26) remained significantly associated.

**Table 5.** Risk factors associated with inpatient readmission

|  | **Bivariate OR** | **CI 95%** | **p value** | **Multivariate OR** | **CI 95%** | **p value** |
| --- | --- | --- | --- | --- | --- | --- |
| Age (years) | 1.01 | (0.965-1.056) | 0.501 |  |  |  |
| Sex (female vs. male) | 0.582 | (0.23-1.47) | 0.25 |  |  |  |
| Comorbidities (present vs. absent) | 1.982 | (0.78-5.04) | 0.144 |  |  |  |
| Hypertension (yes/no) | 2.802 | (1.02-7.63) | 0.039 | 4.99 | (1.54-16.02) | 0.007 |
| Obesity (yes/no) | 3.529 | (0.93-13.47) | 0.085 |  |  |  |
| Diabetes mellitus (yes/no) | 3.529 | (0.93- 13.47) | 0.085 |  |  |  |
| Use of ACE-inhibitors or ARBs (yes/no) | 2.121 | (0.667-6.75) | 0.26 |  |  |  |
| Time since symptom start ($\geq 10$ vs. $< 10$), days | 0.896 | (0.85-0.94) | $< 0.001$ | | | |
| Radiological affection (unilobar vs. others) | 0.478 | (0.16-1.47) | 0.188 |  |  |  |
| Temperature in the emergency ward ($\geq 38$ vs. $< 38$), °C | 10.25 | (2.71-38.66) | 0.003 | 9.03 | (1.89-45.77) | 0.006 |
| Leukocytes ($< 4000$ vs $> 4000$), /μL | 4.49 | (1.47-13.67) | 0.004 | 4.92 | (1.42-17.11) | 0.012 |
| Lymphocytes (800-1200 vs $> 1200$), /μL | 2.817 | (1.1-7.22) | 0.025 | | | |
| LDH ($> 250$ vs $< 250$), U/L | 6.054 | (2.26-16.25) | $< 0.001$ | 6.63 | (2.82-19.26) | 0.001 |
| Eosinophils ($> 200$ vs $\leq 200$), /μL | 0.93 | (0.90-0.96) | 0.234 | | | |
| C-reactive protein (50-100 vs $< 50$), mg/L | 1.948 | (0.67-5.68) | 0.012 | | | |
| Aminotransferases (abnormal vs. normal) | 2.546 | (1.01-6.42) | 0.042 |  |  |  |
| D-dimer (500-1000 vs $< 500$), ng/mL | 0.583 | (0.16-2.10) | 0.569 | | | |
| Azithromycin | 0.187 | (0.07-0.48) | $< 0.001$ | | | |
| Beta-lactams | 2.508 | (1.01-6.26) | 0.043 |  |  |  |
| Non-steroid antiinflammatory drugs | 1.37 | (0.53-3.56) | 0.514 |  |  |  |

ACE: angiotensin-converting enzyme; ARB: angiotensin receptor blockers.

## DISCUSSION

This study describes the experience of applying a management protocol within an emergency Ward to patients with COVID-19, identifying those patients presenting mild cases and a low risk for unfavorable evolution, performing outpatient treatment and follow-up, and avoiding their admission as inpatients. The discharge criteria gathered in this protocol were based on the risk factors for unfavorable development that were described in the available literature,^2,3^ which have been described for hospitalized patients but lack validation for use on outpatients.

The results of our study show that the ambulatory management of patients with low-risk COVID-19 pneumonia is safe, by showing a very low rate of readmissions and no associated mortality. As a reference, both the readmission (6.4%) and mortality (0%) rates of our 314-patient cohort is akin to the FINE I (5.1% readmissions, 0% mortality) and II (8.2% and 0.4%) categories used for community-acquired pneumonia, as published in 1997 by Fine MJ *et al*^4^ on a cohort of 587 and 244 patients respectively. Currently, there are no therapies available with proven efficacy against mild COVID-19, and hence, it is likely that the good prognosis of our patients is more closely related to having been adequately identified than to the therapy administered. Our study shows that it is possible to manage patients with some comorbidities such as hypertension, DM, obesity or asthma as outpatients, whenever such patients are in a good clinical condition. However, the frequency of other conditions (chronic obstructive pulmonary disease, ischemic heart disease, kidney disease or chronic liver disease) in our cohort has been very low, so we are not able to surely say that our results are applicable on such patients.

Secondly, applying a protocol like the one proposed allows avoiding a sizable amount of inpatient admissions for COVID-19 pneumonia (almost 30% in our experience). The lower pressure on the health-care system due to this lower amount of admissions leaves more resources, both material and human, available in the emergency and hospitalization wards. Those resources may be then used for the care of severely ill patients, which could account for a benefit in their clinical development.

Our program has relied on a telephone follow-up after discharge. This follow-up had two goals: on one hand, to monitor the program itself in order to ensure that the proposed management strategy (with no background or prior experience to support it in the available literature) was safe and effective; and on the other hand, to identify and rapidly readmit those patients that showed worsening condition. However, the rate of readmissions has been very low, the severe complications very scarce and mortality has been non-existent in our sample. Moreover, 60% patients that were readmitted had gone back to the emergency ward by their own choice, and the clinical status of the patients reevaluated in the emergency ward was good in general (only one patient showed respiratory failure at the time of readmission). All this suggests that telephone follow-up, once the safety of our protocol is validated, might not be essential, but it might have a role in avoiding a greater number of consultations by providing the patients with a close follow-up and a greater security.

Thirdly, this study allows us to identify the prognostic factors for both good and bad evolution in outpatients with COVID-19 pneumonia. Some of these factors have been previously described for hospitalized patients. Such is the case of hypertension, leukopenia, lymphocytopenia, renal function impairment or LDH and liver enzyme elevations. Some studies like the one done by Liu F *et al*,^5^ or Sun Su *et al*^6^ hint at the fact that a high eosinophil count might be a positive prognostic factor. On that note, the patients in our sample that were not readmitted showed a greater eosinophil median, and no patient with an eosinophil count over 200/µL was readmitted. Some clinical variables seem to be strongly associated with the odds of requiring readmission, such as fever (temperature over 38ºC) at the time of evaluation in the emergency ward. The fact that no patient discharged being on their 10^th^ day of disease progression or later required readmission is noteworthy. This coincides with what has been described about the COVID-19 pathogenesis.^7^ Once the first week has been overcome, patients might enter an inflammatory phase with progressive worsening, (in which case they would meet admission criteria) or start recovering until the fever and the respiratory symptoms are resolved.

There is an important discussion about the effect of angiotensin-converting enzyme (ACE) inhibitors, angiotensin II receptor blockers (ARBs) and nonsteroidal anti-inflammatory drugs (NSAIDs) in patients with COVID-19.^8-12^ In our study, there was no significant relationship to be found between the use of ACE-inhibitors, ARBs or NSAIDs and a bad prognosis. We have not detected that neither the presence nor the absence of certain symptoms (fever, cough, dyspnea, diarrhea) predicts good or bad development, and we also do not observe any significant relationship between the clinical development and the radiological pattern (at least when the injuries affect less than 50% of the lung fields). In our cohort, the D-dimer levels were not correlated with the evolution of the patients, although no patient had a D-dimer level above 1,000 ng/mL (being an exclusion criterion). The association of D-dimer and severity is well described in the literature,^13,14^ but its association with respiratory distress and/or mortality significantly increases with values above said threshold. Therefore, it is possible that its differentiating role is much less relevant for values below 1,000 ng/mL.

Our study has observed a higher rate of beta-lactam use and a lower one for azithromycin in the group that was eventually readmitted. These findings shall be interpreted cautiously, since the study is not designed to evaluate their efficacy and it is highly likely that there are numerous biases affecting them. The therapeutic protocols for COVID-19 have varied very rapidly along the course of this epidemic, and the therapies prescribed may have been strongly affected by subjective criteria depending on each physician, due to the scarce available evidence.

Retrospective analysis of our findings allows us to propose some improvements for our protocol. Slightly elevated D-dimer (yet below 1,000 ng/dL) seems to be of no prognostic value, so they might be withdrawn from the list of minor criteria. To simplify clinical decision-making, the maximum number of minor criteria could be limited to three, which would still include 88.9% of the analyzed patients, but would slightly reduce the risk of readmission. Besides, a short observation period (less than 24 hours) before discharge might be convenient for patients presenting high fever at the time of evaluation in the emergency ward. In any case, new studies are needed to confirm such findings.

## Limitations of this study

This study has some limitations, including having been performed only in one hospital, which may hinder its external validity. Older or more comorbid populations might account for a lower proportion of ambulatorily manageable patients, although some select patients might benefit from it. Another relevant issue is the fact that the study does not include information about the cases that were admitted into hospital upon their first visit, so we do not know if there may have been patients that met discharge criteria but were indeed admitted as inpatients and then eventually required ICU admission or died. However, and as a consequence of the great pressure on the emergency ward of our hospital during the pandemic, the ambulatory follow-up program included almost 50% more patients than the ones analyzed, who did fulfil major admission criteria but were discharged for being clinically stable. Those patients have not been included in the protocol analysis, but they somehow show that the trend at the moment was prioritizing outpatient management whenever possible. It shall be noted that those patients evolved well in general.

Thirdly, the results of the RT-PCR for SARS-CoV-2 is only available in 44% of the cases, and a positive result is only present in 30% of the cohort. We consider this to be a consequence of two factors. The protocols from the valid health authority at each moment have changed widely during the pandemic. During most of the study period, performing the test was only authorized in dubious cases, and hence the diagnosis of typical cases was based on clinical, radiological and laboratory findings. Besides, 29% of the patients on whom PCR was performed have a negative result. We do not believe this fact to impair the reliability of our results, since PCR sensitivity in our institution was found to be 65-70%, only PCR-negative patients with high clinical suspicion were included in the study, and positive predictive value for COVID-19 diagnosis in Madrid, during the peak of the pandemic was remarkably high. In addition, there were more tested patients among those who were readmitted. The explanation could be that chances of be tested were higher for them, as they were finally admitted into hospital for a clinical deterioration.

Finally, we have detected a great difference between groups regarding the prescribed treatment. This might be a problem in the setting of highly effective treatments, but the evidence available at the moment does not suggest that beta-lactams or azithromycin have a relevant therapeutic role in COVID-19 pneumonia.^15-17^

## CONCLUSIONS

In the setting of a great overload of the healthcare system and risk for collapse, the identification of low-risk patients for their ambulatory management is fundamental in order to free hospital resources that then may be destined for more severe patients in greater need. The outpatient management of patients with low-risk COVID-19 pneumonia is safe, whenever they are adequately selected. The management protocol presented in this study has allowed avoiding 30% of the admissions for COVID-19 pneumonia in our hospital, with a very low readmission rate and non-existing mortality. Hypertension, high fever at the time of evaluation, leukopenia and increased LDH were associated to a higher risk for readmission, while a high eosinophil count and a longer time since the start of symptoms were associated with low risk. In addition, mild elevation of D-dimer did not seem to have prognostic relevance.

**Figure 1.**
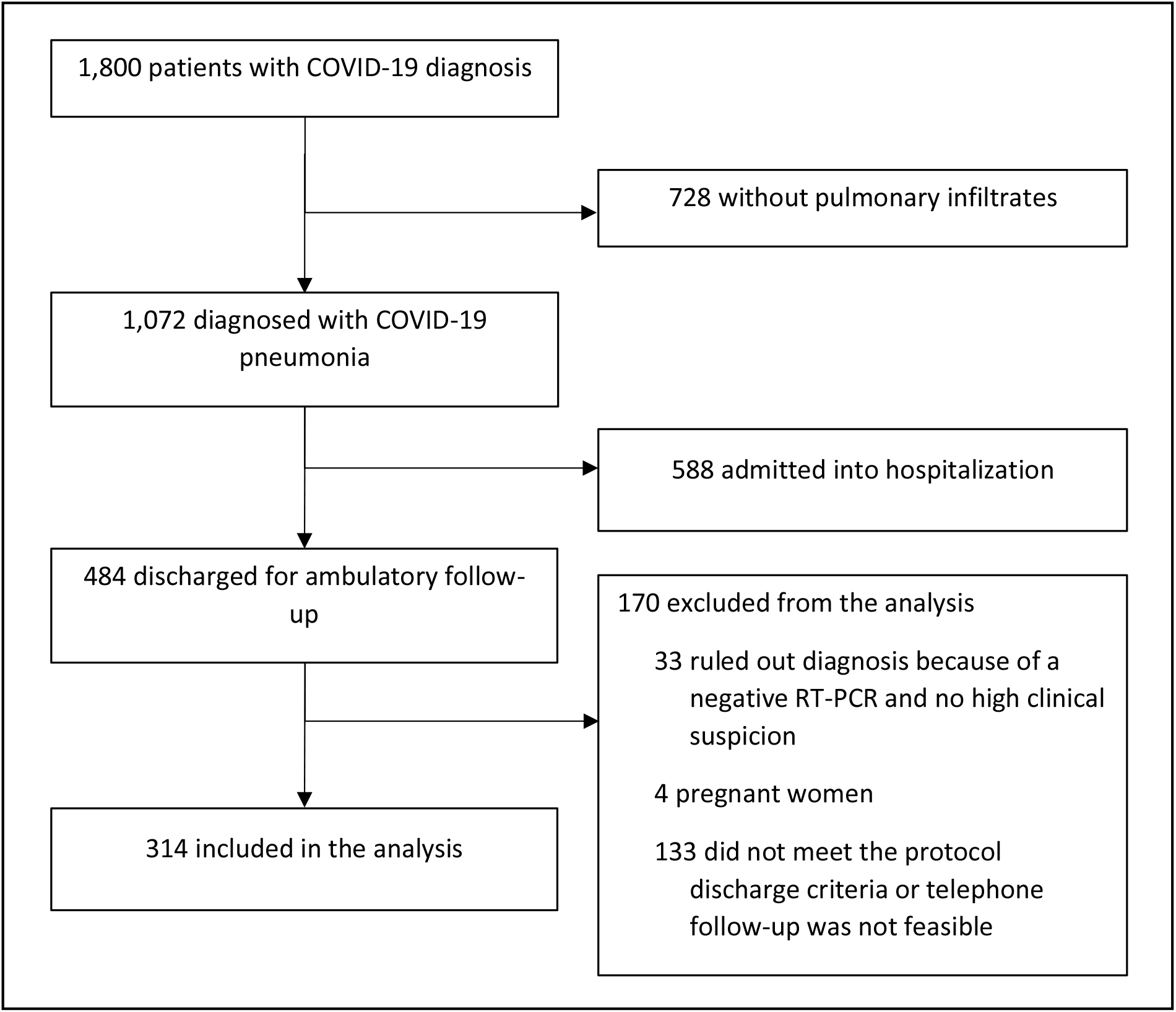
Population included in the analysis.

## Data Availability

The datasets generated during and/or analysed during the current study are available from the corresponding author on reasonable request

